# Improving Post Kidney Transplant Related Outcomes through Video Based Educational Platforms

**DOI:** 10.1101/2025.01.29.25321359

**Authors:** Jennifer Byrns, Jason Bodner, Michael Watson, Tresa Conca, John Roberts

## Abstract

**PURPOSE:** Kidney transplant recipients’ medical knowledge and comprehension of transplant care are critical to successful long-term outcomes. While transplant centers focus on pre-transplant patient education, post-transplant education receives less emphasis. At Duke University Hospital, an online digital library of short patient-centric videos was created to provide transplant education. The purpose of this study was to evaluate the impact of these videos on improving patient outcomes and patient perceived ability to effectively manage their own care.

**METHODS:** This was a prospective, single center study, evaluating the impact of education via patient-centric videos on medical knowledge, medication adherence, hospital readmissions, and patient satisfaction. Patients were randomized to receive education via online videos versus standard of care (in person and written education). Ten videos were created to cover the entire transplant process. A survey was conducted to assess outcomes. Descriptive statistics were used to compare groups with absolute numbers (%) and means (SD).

**RESULTS:** There were a total of 83 patients enrolled. Response rates to the surveys was 61.4%. Patients who completed the survey were evaluated. Twenty seven patients were included in the video group and there were 24 control patients. The video group expressed extreme satisfaction with the videos. All patients reported that they would recommend the videos to other patients, and 84% stated that they would return to watch the videos to gain additional transplant knowledge. Patients in the video group reported confidence in their ability and the medical team’s ability to manage their health. The assessment of transplant knowledge was similar between groups, but limited by video completion rates and timing of survey completion. Readmission rates were no different.

**CONCLUSIONS:** Transplant education provided via video based format helped to improve patients’ confidence in their ability to maintain their own self-care. The videos were well received and helped give patients a method to learn about transplant. These videos are now used institution wide to help improve patient outcomes

## INTRODUCTION

Kidney transplantation provides patients with end stage kidney disease the opportunity to have an improved quality of life and overall longer term survival. The benefit of longer term survival is directly related to function and longevity of the kidney allograft. In order to preserve allograft function, patients are required to take complex immunosuppressant regimens and adhere to lifestyle modifications to prevent graft rejection and avoid graft loss. The complexity of these regimens can be difficult for patients to understand and follow post-transplant.

Health literacy is one factor that has been positively associated with adherence to medical regimens. Interventions to improve literacy have had great impact in those with lower income and/or racial-ethnic minority patients.^1^ Medication compliance is also directly affected by health literacy and social support.^2, 3^ The prevalence of non-adherence to medications in kidney recipients has been quoted at 35.6 cases per 100 people per year, monitoring vital signs at 20.9 cases per 100 people per year, and diet at 22.6 cases per 100 people per year.^4^

Education is a means to help improve health literacy and assist kidney transplant recipients navigating the complexity of the post-transplant course. Current platforms for education include verbal education performed by multidisciplinary team members in the inpatient and outpatient setting as well as written materials provided at discharge. These teaching mechanisms are often standardized and not modified based on varying degrees of literacy. The comprehension of these materials are often not assessed in the post-transplant setting. Newer forms of educational materials such as video based platforms have shown success in the pre-transplant setting but more literature is needed for developing videos for education post-transplant.^5^

At our institution, a study was conducted to develop and provide animated short education videos to patients during the early post-transplant course. These videos were constructed to address and educate on various areas of transplant care that the patients could reference at any point during the study period. The purpose of this study was to evaluate the impact of patient centric video-based education on improving patient outcomes and the perceived ability to effectively manage their own care.

## METHODS

### Study Design

This was a prospective, open-label, randomized controlled study of kidney transplant patients who were transplanted at Duke University Hospital between July 1, 2019 to August 1, 2021. The study protocol was approved by the Duke University Hospital Institutional Review Board and is in alignment with the ethical guidelines of Helsinki Declaration.

Patients were randomized 1:1 into two groups: a video based platform with standard of care education provided by the multidisciplinary transplant team both in the inpatient and outpatient setting (verbal and written education) versus standard of care only education.

Patients and/or caregivers were consented for enrollment in this study. All participants indicated on the consent form how they would like to receive the videos if randomized into the video group (text message to phone, links sent via the online patient portal, email, or watching on tablet computer in clinic).

### Inclusion and Exclusion Criteria

Patients were included in this study if they completed the consent process within 7 days of receiving a kidney transplant, were 18 years of age or older, and had the capacity to give legal written consent.

Patients were excluded if they had severe vision or auditory impairments rending them unable to view or hear videos at a close range, patients unable to understand the English language, or those with delirium, dementia, or other cognitive impairments.

### Objectives

The primary objective was to assess the impact of patient centric video curriculum on transplant medical knowledge.

Secondary objectives included the impact of the patient centric video curriculum on perceptions of self-management efficacy post-transplant, satisfaction with the videos, hospital readmissions, and medication errors noted during pill box reviews in the outpatient setting.

### Video Development and Implementation

The video curriculum Content was developed through several focus group sessions with a multidisciplinary group of full-time transplant team members (transplant pharmacists, nurse practitioners, physician assistants, nurse coordinators, surgeons, and nephrologists) who actively manage kidney transplant patients at our transplant center and are familiar with post-transplant education. As a result of these focus group sessions, we identified the core topics to be included in the video curriculum. This included the following: a welcome to kidney transplant video educating patients on what kidney transplant is and who their transplant medical team members were; complications and expectations after kidney transplant; fluid and blood pressure management after kidney transplant; de-mystifying kidney lab work which educated on what lab values represent and how the team interprets them; diabetes after transplant (2 part series) with types of diabetes, medications, health tips on diabetes control and healthy lifestyle, how to monitor/test blood glucose, how to treat hypo/hyperglycemia; recovery and questions after transplant with answers to commonly asked questions; and transplant medications (3 part series) with teaching on commonly used medications post-transplant, and how to manage these medications.

Video production was standardized and designed to boost viewer engagement. The narration for each video was scripted ahead of time by the authors, and reviewed, edited, and approved by our institutional Patient Family Education Governance Council to maximize patient-centered language. We produced ten videos using the Powtoons® production platform. All videos were produced according to published guidelines shown to boost viewer engagement.^6^ The videos were short (4-7 minutes in duration), used animated cartoons and figures with minimal text, and narrated using a conversational tone.

The videos were distributed to patients via SMS text message to phone numbers provided during consent, message via Duke MyChart or through email, or watching on a tablet computer in clinic which had the videos preloaded to view during transplant visits.

### Survey Development and Implementation

A survey was created to assess knowledge, self-management, medication adherence, and satisfaction. The survey was provided in several sections including the Patient Activation Measure (PAM 10), a validated instrument that measures patients’ perceptions of knowledge, skills, and confidence in self-management.^7^ The survey was divided into blocks to enhance response efficiency.

The first patient activation measure (PAM) block assessed the ability and confidence to manage one’s own care. This block consisted of 10 Likert scale questions including: I am confident that I can maintain lifestyle changes like eating right and exercising, even during times of stress; I am confident I can figure out solutions when new problems arise with my health; I know how to prevent problems with my health; I have been able to maintain (keep up with) lifestyle changes, like eating right or exercising; I am confident that I can follow through on medical treatments I may need to do at home; I am confident that I can tell a doctor concerns I have even when he or she does not ask; I am confident that I can tell whether I need to go to the doctor or whether I can take care of a health problem myself; I know what each of my prescribed medications do; taking an active role in my own health care is the most important thing that affects my health; when all is said and done, I am the person who is responsible for taking care of my health.

The second block of questions consisted of those related to the patient’s ability to maintain adherence to their medication regimen. This section asked questions about self-reports on adherence over a one week period, obtaining refills on time, and concerns about adverse effects of medications.

The third block of questions were split into 2 PAM based blocks focusing on the ability and the confidence of the patient to manage one’s own care and find tools to help and the confidence in medical team to help them in their care. Five likert scale questions were asked in the first half of the patient management reflection block including: If I am in trouble, I can reach out to my team for solution; I know how to get help for unforeseen medication situations; I can usually handle whatever comes my way; I am confident that I could deal with unexpected medical events; and I can manage difficult problems if I try hard enough. Both groups responded with having the resources required to manage their own care. The distribution of these responses were similar between groups although a few more patients in the video group responded with neither agree nor disagree. The second half of the block focusing on the patient perceived confidence in the medical team consisted of 5 likert scale questions including: how confident are you that you can do all the things necessary to manage your kidney transplant on a regular basis; how confident are you that you can judge when you need to contact your transplant team for health problems; how confident are you that you can ask your doctor things about your transplant or medications that concern you; how confident are you that you can discuss openly with your doctor any personal problems related to your transplant or medications; and how confident are you that you can work out differences with your doctor when they arise.

A transplant knowledge block of questions were administered in the fourth block (Supplemental Table 1). These questions ranged from understanding who to contact for medical questions or concerns, determination of common transplant labs for kidney transplant patients, understanding rejection, indications for transplant medications and common adverse drug effects, how to monitor vitals and the frequency, common post-operative complications, who transplant team members are and their role, and along with various medical scenarios and the best way to handle them.

The final block of likert type questions was only provided to the video group. Questions focused on satisfaction with the videos and the likelihood of recommending the video to others and/or returning to view the videos in the future.

The survey was sent to patients after 3 months post-transplant, allowing a reasonable amount of time to review the videos outside of the index transplant admission. Study investigators reached out to patients who had not completed the survey at the 6 month mark after consent. To improve response rates and compensate for time, patients who completed the survey were provided with a Visa gift card valued at $50.00 (USD).

### Statistical Analysis

Descriptive statistics were used to analyze the primary and secondary objectives of this study. Continuous variables were repeated using mean (standard deviation [SD]) and median (interquartile range [IQR]) where appropriate. Categorical variables were reported using frequencies and percentages. The two groups were compared using student’s t-test or Wilcoxon rank sum tests for continuous data and fisher’s exact test or chi-square tests for categorical variables. Significance of all tests was assumed if the p-value ≤ 0.05. All statistical analysis was conducted using GraphPad Prism® version 10.0.0.

## RESULTS

83 patients completed the consent to participate. 79 patients completed the study during follow-up: 3 patients withdrawing due to preference and 1 withdrew due to graft loss. A total of 51 patients completed the survey. The video group consisted of 27 patients and the control group had 24 patients.

Baseline demographics were collected on all patients (see Table 1). The 2 groups were similar except for baseline education level. There were more patients with advanced degrees (bachelor’s, master’s, or other degrees) in the control group compared to the video group (p=0.05). Occupation was also collected as a part of this study with more patients in the video group being in business management versus the control group with education and training.

**Table 1.**
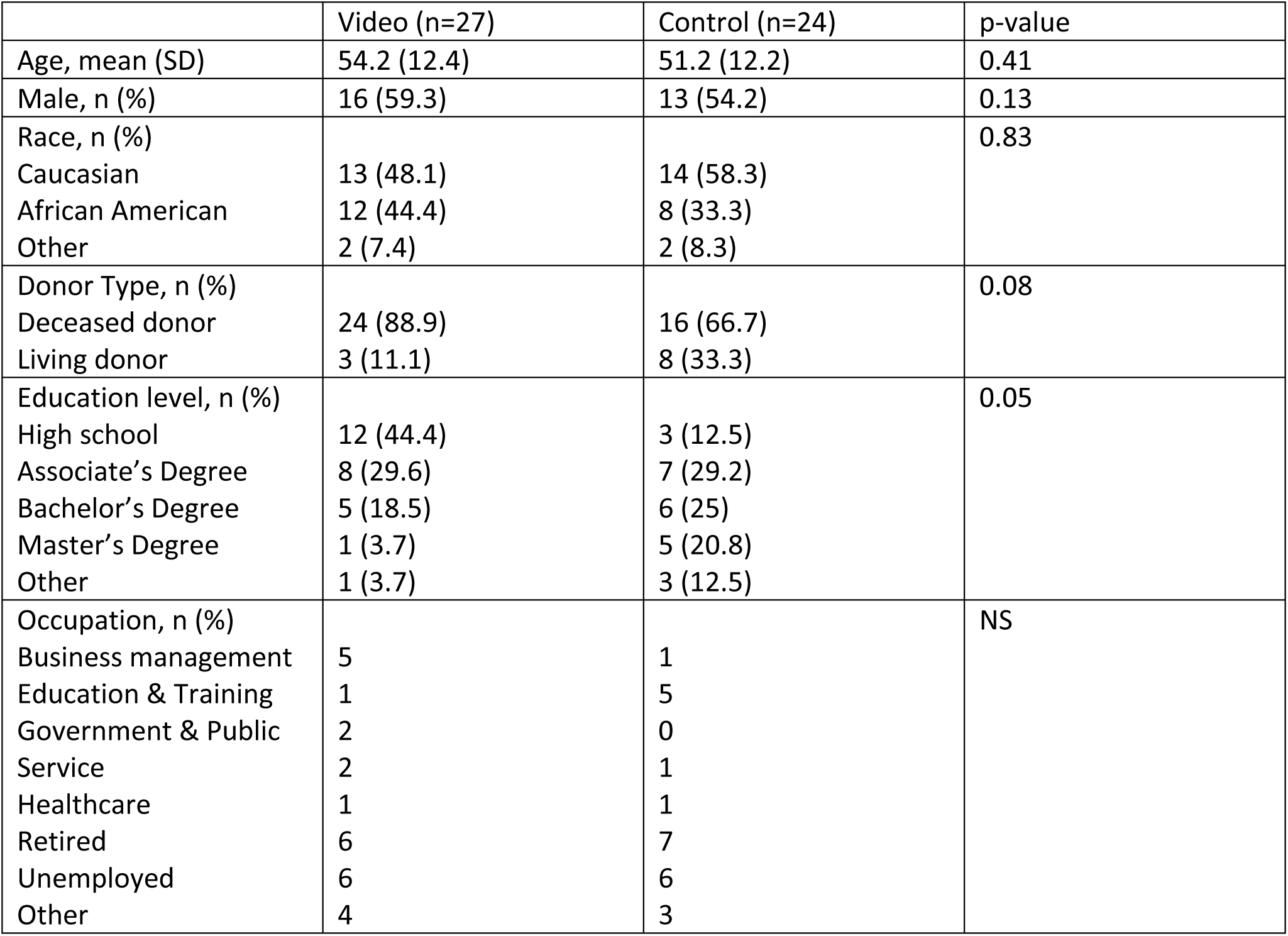
Demographics.

We analyzed survey results according to each block. The first block consisting of patient activation and self-management questions found that both groups equally agreed >95% of the time that they were confident about their abilities to care for them own health (Figure 1). The video group had a wider distribution of responses to this measure. The second block examining self-reported medication adherence, revealed that in each group there was a high likelihood of medication adherence with the 88.9% of the video group reporting never missing a dose of medication and 11% of patients reporting 1 missed dose in a week (Table 2). The control group reported 100% adherence. The video group was more likely to report concerns about their transplant medication side effects (66.7% reported being concerned most of the time or always when compared to 8% in the control group).

**Figure 1.**
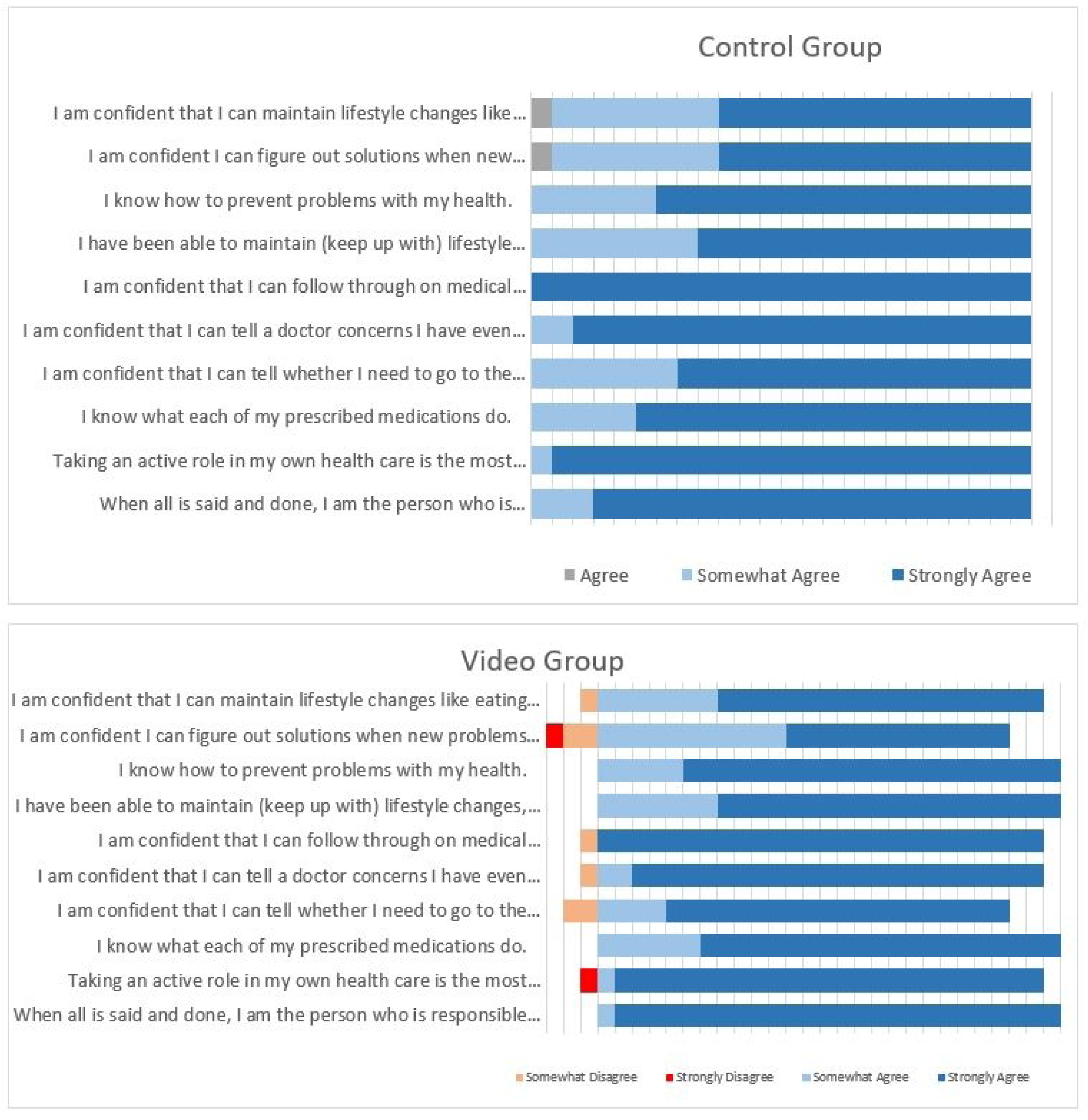
PAM Block 1 - Ability/Confidence to manage

**Table 2.** Medication Adherence Block.

| How often do you miss doses of medications over the course of a week? |  |  |
| --- | --- | --- |
|  | Video (n=27) | Control (n=24) |
| Everyday | 0 | 0 |
| 4-6 times a week | 0 | 0 |
| 2-3 times a week | 0 | 0 |
| Once a week | 2 | 0 |
| Never | 25 | 24 |
| I run out of medications because I don't get refills on time |  |  |
| Always | 0 | 0 |
| Most of the time | 0 | 0 |
| About half of the time | 1 | 0 |
| Sometimes | 3 | 0 |
| Never | 23 | 24 |
| I worry about my transplant medication side effects |  |  |
| Always | 9 | 2 |
| Most of the time | 9 | 0 |
| About half of the time | 1 | 4 |
| Sometimes | 6 | 6 |
| Never | 10 | 12 |

The third block, split into 2 PAM based questions, found that both groups responded equally to having the resources required to manage their own care. The distribution of these responses were similar between groups although a few more patients in the video group responded with neither agree nor disagree (Figure 2).The second section of this block examined the patient’s confidence in the medical team with scores out of 100% assigned based on the level of confidence. Both groups responded with an overall 96% confidence in their ability to manage their transplant and discuss issues with the medical team.

**Figure 2.**
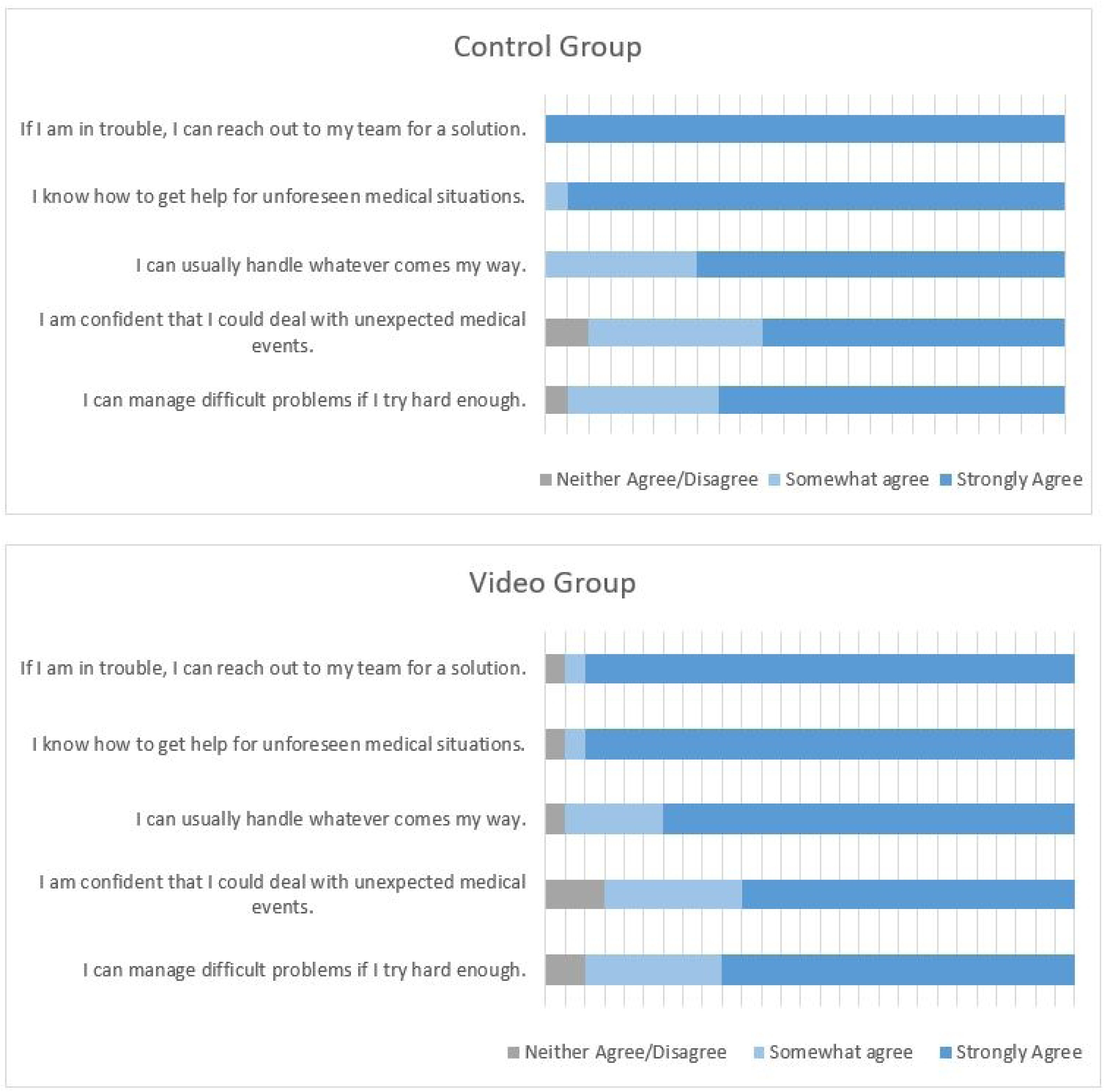
Self-Efficacy and Resiliency: Ability/Confidence

The transplant knowledge block, which was scored based on the number of correct answers, found that 2, the control group provided more correct answers than the video group on both block 1 and 2 (block 1 median 93.8% vs 87%, control vs video group respectively; block 2 median 81.8% vs 74.1%). Of note, only 22 patients in the control group were analyzed for this block due to lack of response on this section by 2 patients.

The final block of questions related to video satisfaction determined that, patients in the video group were extremely satisfied and reported being very likely to recommend the videos to others (Table 3). Their preferred form for receiving the videos was via text message. Patients also self-reported when they watched the videos for this study. Three patients out of the 27 (11.1%) reported having not watched the videos, 2 patients (7.4%) watched the videos >60 days after enrollment, the other 22 patients (81.4%) watched the videos within the first 2 weeks after enrollment.

**Table 3.** Video Satisfaction.

|  | Video (n=25) |
| --- | --- |
| Overall, how satisfied or dissatisfied were you with the transplant videos? n (%) |  |
| Extremely satisfied | 18 (72) |
| Somewhat satisfied | 5 (20) |
| Neither | 2 (8) |
| Somewhat dissatisfied | 0 |
| Extremely dissatisfied | 0 |
| On a scale from 0-10, based on your experience, how likely are you to recommend the transplant videos to another patient? n (%) |  |
| 10 | 17 (68) |
| 9 | 3 (12) |
| 8 | 3 (12) |
| 7 | 0 |
| 6 | 1 (4) |
| 5 | 1 (4) |
| How likely would you be to return to watch the transplant videos? n (%) |  |
| Extremely satisfied | 12 (48) |
| Somewhat satisfied | 9 (36) |
| Neither | 4 (16) |
| Somewhat dissatisfied | 0 |
| Extremely dissatisfied | 0 |
| How easy was it to watch the videos? (24 responses) n (%) |  |
| Extremely easy | 19 (79.2) |
| Somewhat easy | 3 (12.5) |
| Neither | 2 (8.3) |
| Somewhat difficult | 0 |
| Extremely difficult | 0 |
| What was your preferred method for watching the videos? (25 responses) n (%) |  |
| Text message links | 17 (68) |
| Through mychart | 7 (28) |
| The tablet (ipad) in clinic | 1 (4) |
| How much more time would you like to spend watching videos like this in the future? (24 responses) n (%) |  |
| A great deal | 1 (4.2) |
| A lot | 4 (16.7) |
| A moderate amount | 12 (50) |
| A little | 6 (25) |
| None at all | 1 (4.2) |

Survey completion rates were low in both cohorts. Study investigators contacted patients via phone, the online patient portal, and by text message to remind them to complete the surveys. Only 31 out of the 83 enrolled patients completed the survey by 6 months. An additional 20 patients completed the survey after 1-2 reminders to complete these before the end of the first year post-transplant.

A separate analysis was performed on patient outcomes between groups on readmission rates (including emergency department (ED) visits) and pill box errors seen in clinic by the transplant pharmacist. Nine patients in the video group were readmitted or visited the ED within the first year post-transplant versus 11 patients in the control group. Medication errors were found in 5 (17.9%) of patients in the video group and 5 (20.8%) of the control group.

## Discussion

This study examined the creation and use of a video based curriculum to provide education to kidney transplant recipients in the post-transplant setting. The videos were designed to target focused areas during the post-transplant course that patients will encounter, such as complications and expectations after kidney transplant, fluid and blood pressure management, common lab tests, diabetes, recovery post-transplant, and transplant medications. Patients in the video group were overall satisfied with the content and reported they were easy to watch. They were also very likely to recommend others to watch the videos. Both groups displayed a comparable confidence in their abilities to manage their own healthcare or find tools to assist them. When assessing the knowledge gained by the videos, there did not appear to be a difference between the 2 groups despite having an additional resource for education.

At this time, there is limited literature analyzing the effectiveness of video based education in the post kidney transplant setting. Most of the literature with video-based platforms is focused on the pre-transplant phase with educational interventions targeting deceased donor organ acceptance, living donation, and knowledge about transplant^8–10^ Providing education in this phase of care has shown positive results with improving knowledge. Our study attempted to show an improvement in knowledge gained in the post-transplant phase. Although the knowledge assessment was similar between groups, it did help the study investigators identify areas that can be improved in our transplant education overall. These topics include understanding rejection, dietary choices, adverse drug effects, and diabetes management.

Post-transplant educational videos allow for another modality to provide education to patients. One of the key measures for providing education to a diverse patient cohort is understanding how patients’ best receive information (ie. visual, auditory, kinesthetic, etc).^11^ Assessing this during the transplant phase can help gear the delivery of the education properly. Videos that are animated and visually appealing can provide a tool to engage active learning. At our institution we are currently using these videos to complement the standard written and verbal communication we give in the hospital and during clinic visits. Patients that display a lack of knowledge in an area are given these videos to view at home to help improve their understanding. We also provide all of the video links via QR codes in the discharge paperwork at the index transplant surgery discharge and during clinic visits after ambulatory patient encounters.

There were several limitations to this study. The groups were not balanced based on educational background. There were more patients with advanced degrees in the control group which may have affected health literacy, knowledge, and measures of perceived self-efficacy. Survey completion rate were also low and there was a delay in completing the surveys with several patients completing it after 6 months post-transplant. Furthermore, not all patients completed the full survey. Patients were included in the analysis if they completed at least 80% of the questions. In the video group, participation in viewing the videos in their entirety was self-reported but not validated through a tracking mechanism. Patients could also view all the videos in succession which may be overwhelming with the amount of education provided at once. Pushing out the videos to be viewed at differing times post-transplant may help improve knowledge retention. This study was also conducted during the height of the COVID pandemic which likely affected study participation and completion rates. This also affected the time it took to analyze and publish the results of this study.

## Conclusion

Video based educational platforms used in the post-transplant setting can be a tool to enhance learning on complex topics. Developing videos that are engaging, easily accessible, and viewable at any time can help complement current education given after transplant. These videos can help empower patients to gain knowledge at their own pace and can be provided by the transplant team at various times when patient needs are identified. Overall post-transplant video education is well received by kidney transplant patients.

## Data Availability

All data is available, without restriction, for review if needed.

---

**Supplemental Table 1:** Transplant Knowledge Base Questions (multiple choice)

| Transplant Knowledge Block 1 |  |
| --- | --- |
| After transplant, if you started feeling sick and had a fever, what would you do next? | A. Take Tylenol and see if the fever goes away<br>B. Temporarily stop taking Prograf<br>C. Temporarily stop taking diuretics (water pills)<br>**D. Call transplant coordinator or go to urgent care/hospital** |
| If you started vomiting so much that you couldn't hold down your kidney transplant medications, what would you do next? | A. Try to drink lots of water to stay hydrated<br>B. Take some pepto-bismol<br>C. Rest at home, give it two to three days to see if it gets better<br>**D. Call transplant coordinator or go to urgent care/hospital** |
| Ms Jones just had a kidney transplant one month ago. When looking at her labs, all of her creatinine tests show that the creatinine is going up every time it is checked. This means her kidney function is... | A. Getting better<br>**B. Getting worse**<br>C. Staying about the same |
| How can you tell whether someone has kidney disease or not? | A. Seeing if they have swelling in the feet<br>B. Looking at an X-ray of their lungs<br>**C. Looking at blood tests including serum creatinine**<br>D. Looking at the color of their urine |
| Right after transplant, you are asked to taper (reduce) your prednisone by 5mg (1 pill) each week. Which medicine below is prednisone? | Provided with 4 pictures of pills (tacrolimus, prednisone, mycophenolate capsules, mycophenolate tablets) |
| When you get transplanted, we start you on immunosuppression medications. What is the purpose of these medicines? | **A. Prevent kidney rejection**<br>B. Prevent infections<br>C. Control blood pressure<br>D. Remove fluid by making you pee more |
| Which of the following would be a good reason to call your transplant coordinator? | A. Nurse from family doctor called and wanted a call back<br>B. Family member having symptoms<br>C. Need payment for duke parking<br>**D. Diarrhea more than 8 times a day and feeling dizzy** |
| If you had questions about what happened during your transplant surgery, who is the best person to ask? | A. Transplant nephrologist<br>B. Transplant coordinator<br>C. Transplant nutritionist<br>D. Scheduled at front desk<br>**E. Transplant Surgeon** |
| If you need standing lab orders (for blood work) faxed to the lab near where you live, who is the best person to contact | A. Transplant pharmacist<br>**B. Transplant coordinator**<br>C. Transplant social worker<br>D. Transplant surgeon |
| The best way to monitor for fluid build up on your body is.. | **A. Measuring weight and checking legs for swelling each day**<br>B. Puffiness around the eyes<br>C. Seeing if you feel short of breath or not<br>D. Feeling bloated or not |
| When recovering from kidney transplant surgery, if you measured a very low blood pressure (85/55), what should you do next? | A. Take all medications and call coordinator<br>**B. Hold (don't take) blood pressure medications or fluid pills and call my coordinator**<br>C. Go for a long walk outside<br>D. Drink a glass of water immediately |
| After kidney transplant, if your legs started swelling AND you were gaining weight (up 10 pounds in 5 days), what would you do next? | A. Add salt to the food I eat<br>B. Elevate my legs each night<br>C. Drink 8 glasses of water each day<br>**D. Call my coordinator to let the team know** |
| Which of the following statements is NOT true? | A. People can get diabetes for the first time shortly after kidney transplant<br>B. Transplant pills like prednisone can lead to high blood sugars<br>C. High blood sugars can make it hard for the wound to heal<br>**D. Having sugars above 300 mg/dl is ok and causes no harm to your body after transplant** |
| Which of these foods will NOT raise your blood sugar as much as the others? | A. Sweet tea<br>B. Biscuits and Gravy<br>C. French fries<br>**D. 8 oz sirloin steak** |
| Which of these foods will NOT raise your blood sugar as much as the others? | **A. Baked chicken**<br>B. Ice cream<br>C. Mashed potatoes<br>D. Chocolate chip cookies |
| Which of these foods will NOT raise your blood sugar as much as the others? | **A. Roasted carrots**<br>B. Regular coke<br>C. M&Ms<br>D. White rice |
| **Transplant Knowledge Block 2** |  |
| What is kidney rejection? | A. My immune system attacking my transplanted kidney<br>B. My transplanted kidney<br>C. My old kidneys stop working |
| What are possible signs of rejection? Select all that apply | A. Numbness in hands or feet<br>**B. Fever greater than 100.5**<br>**C. Drop in urine output**<br>D. Headache that lasts more than 1 day<br>**E. General flu-symptoms, just feeling "Bad"** |
| What are possible problems that can happen after I receive my kidney transplant? | A. Infections<br>B. Getting diabetes for the first time<br>C. Having high blood pressure |
|  | D. Surgery wound infections<br>**E. All of the above** |
| Who is part of my post-kidney transplant team?<br>Click the correct team members? | **A. Post-transplant coordinator**<br>**B. Nephrologist**<br>**C. Surgeon**<br>D. Primary care provider<br>**E. Pharmacist**<br>**F. Dietitian** |
| Why is it important for me to make healthy food choices? Click all the options that are correct. | **A. Healthy foods help give my body energy to heal**<br>**B. Foods high in sugar can increase my risk for obesity and developing diabetes**<br>**C. Health foods help build muscle and improve strength**<br>D. Food with protein can hurt my kidney |
| What is considered a high blood pressure right after transplant? | **A. BP 160/90**<br>B. BP 134/80<br>C. BP 122/72 |
| Select the correct options for when to contact coordinator now versus waiting until your next transplant doctor's appointment | Fever >100.5 – Coordinator<br>Incision is draining or bleeding – Coordinator<br>Ran out of medication – Coordinator<br>Traveling to another state in a few weeks to visit family – Next appt<br>Difficulty urinating – Coordinator<br>Need payment for duke parking – Next appt<br>Nurse from family doctor called and wanted to call back – Next appt<br>Hair starting to fall out – Next appt |
| How many times should you measure and record the following after transplant? | Blood pressure – two times a day<br>Temperature – two times a day<br>Weight – once a day |
| Select the medications that are used to prevent rejection | **A. Prograf (tacrolimus) and Envarsus (tacrolimus-XR)**<br>B. Bactrim (sulfamethoxazole/trimethoprim)<br>**C. Cellcept (mycophenolate)**<br>**D. Prednisone** |
| Which transplant medication needs a blood test to determine the correct dose? This is the one we ask you to hold before lab draws in the morning | Provided pictures of tacrolimus and tacrolimus-XR; mycophenolate, prednisone |
| Select the side effects that occur with each transplant medication | Mood changes – prednisone<br>Increased appetite – prednisone<br>Nausea/Vomiting – mycophenolate<br>Tremors – tacrolimus<br>Diarrhea – mycophenolate<br>Headaches – tacrolimus<br>High blood sugars – prednisone |
| Match the medication with the infection it prevents | Bactrim (sulfamethoxazole/trimethoprim) – prevents bacterial infection<br>Valcyte (valganciclovir) – prevents viral infections<br>Zovirax (acyclovir) – prevents viral infections |
| What is the BK virus and why is it bad? | **A. It affects mostly kidney transplant patients and if the levels get high enough, it can hurt the kidney**<br>B. It can affect any patient, and causes brain problems<br>C. It's a common virus that affects transplant patients that can give you diarrhea |
| You are asked to fill your pill box with mycophenolate 1000mg twice daily. Each capsule strength is 250 mg. Please touch the individual boxes where you would place your mycophenolate capsules | Interactive selection for AM and PM boxes |
| What is the maximum amount of time you can miss a dose of medication and still be able to take it? | A. 15 minutes<br>**B. 4 hours**<br>C. 12 hours<br>D. 24 hours |
| Select the appropriate times to contact your coordinator with medication questions. | **A. Confirming how to take your medications (ie. number of tablets and at what time)**<br>**B. Upcoming change in prescription insurance**<br>**C. No refills left on prescription medication**<br>**D. Want to start a new over the counter medication**<br>**E. Will be running out of medication at end of week and need pharmacy to refill prescription** |
| What is the best method to determine the directions on how to take your medications?<br>Select the graphic that matches the best option? | Provided with picture of pill bottle and of **medication grid** |
| Why do some people need to check their blood sugars after transplant? | **A. Transplant medication can increase blood sugars**<br>B. Having a new kidney can increase blood sugars<br>C. Only people who had blood sugar issues before transplant need to check after transplant |
| Which picture best shows how to check your blood sugar? | Provided with picture of glucometer, insulin pen, and blood pressure monitor |
| Which locations on the body can you give insulin shots? Select all that apply. | **A. Abdomen (stomach)**<br>**B. Back of arm**<br>**C. Thigh**<br>D. Calf<br>E. Hands |
| Which reading is considered a low blood sugar and should be treated? | **A. Blood sugar of 65**<br>B. Blood sugar of 142<br>C. Blood sugar of 93 |
| High blood sugars over time can damage which organs? Select all that apply | **A. Kidney**<br>**B. Nervous System**<br>**C. Heart**<br>**D. Eyes**<br>**E. Digestive system** |

## References

1. Miller TA. Health literacy and adherence to medical treatment in chronic and acute illness: a meta-analysis. Patient Educ Couns 2016;99(7):1079–1086. doi:10.1016/j.pec.2016.01.020

2. Gokoel S, Gombert-Handoko K, Zwart T, van der Boog P, Moe D, de Fijter J. Medication non-adherence after kidney transplantation: a critical appraisal and systematic review. Transplant Reviews 2020;34(1):100511. doi: 10.1016/j.trre.2019.100511

3. Demian MN, Shapiro RJ, Thornton WL. An observational study of health literacy and medication adherence in adult kidney transplant recipients. Clin Kidney J. 2016;9(6):858–865. Doi: 10.1093/ckj/sfw076

4. Kidney Disease: Improving Global Outcomes (KDIGO) Transplant Work Group. KDIGO clinical practice guideline for the care of kidney transplant recipients. American Journal of Transplantation 2009;9(Suppl 3):S1–S157.

5. Rosaasen N, Mainra R, Kukha-Bryson A, Nhin V, Trivdei P, Shoer A, Wilson J, Padmanabh R, Mansell H. Development of a patient centered video series to improve education before kidney transplantation. Patient Educ Couns 2018;101(9):1624–1629.

6. Guo PJ, Kim J, Rubin R. How video production affects student engagement: an empirical study of MOOC videos. In proceedings of the first ACM conference on learning@ scale conference 2014:41–50. Doi 10.1145/2556325.2566239

7. Hibbard JH, Stockard J, Mahoney ER, Tusler M. Development of the Patient Activation Measure (PAM): conceptualizing and measuring activation in patients and consumers. Health Serv Res. 2004 Aug;39(4 Pt 1):1005–26. doi: 10.1111/j.1475-6773.2004.00269

8. Kayler LK, Dolph BA, Cleveland CN, Keller MM, Feeley TH. Educational Animations to Inform Transplant Candidates About Deceased Donor Kidney Options: An Efficacy Randomized Trial. Transplantation Direct 6(7):p e575, July 2020. | DOI: 10.1097/TXD.0000000000001026

9. Kayler LK, Seibert RE, Dolph BA, et al. Video education to facilitate patient outreach about living kidney donation: a proof of concept. Clin Transplant. 2021; 35(12):e14477. 10.1111/ctr.14477

10. Mansell H, Rosaasen N, Wichart J, Mainra R, Shoker A, Hoffert M, Blackburn D, Liu J, Groot B, Trivedi P, Willenborg E, Amararajan M, Wu H, Afful A. A Randomized Controlled Trial of a Pretransplant Educational Intervention in Kidney Patients. Transplantation Direct 7(10):p e753, October 2021. | DOI: 10.1097/TXD.0000000000001202

11. Anouk J.L. Muijsenberg, Sarah Houben-Wilke, Yuqin Zeng, Martijn A. Spruit, Daisy J.A. Janssen. Methods to assess adults’ learning styles and factors affecting learning in health education: A scoping review. Patient Education and Counseling 2023;107:e107588. 10.1016/j.pec.2022.107588.

